# Retrospective Evaluation of an AI System to Classify Negative Musculoskeletal Trauma Radiographs

**DOI:** 10.1101/2025.11.27.25341142

**Authors:** Steffen Czolbe, Jonas Christophersen, Rikke Bachmann, Janitha Mudannayake, Andreas Nexmann, Fabizio Perria, Nikolaus Knauer, Pavel Lisouski, Michael Lundemann

**Affiliations:** Radiobotics ApS

## Abstract

**Study objectives:** To evaluate an AI system for musculoskeletal (MSK) radiography in confidently identifying examinations *without* injury-related pathologies to support AI-guided discharge of patients without traumatic findings.

**Methods:** We retrospectively sampled radiographic examinations, including one or more radiographs and a radiological report, of suspected MSK trauma from *>*1,000 clinical sites in two countries. Medically trained professionals independently classified all exams. When disagreements between the classification and the original radiological report arose, adjudication was performed by a third professional. Annotators were also asked to record their confidence level of each classification. The AI system analyzed all exams and assigned them to the categories *AI Positive, AI Negative*, and *AI Negative (very high confidence)*. Performance of the AI system was assessed using error rate, false negative rate (FNR), and qualitative review of misclassified exams.

**Results:** A total of 2,962 exams were included. The AI classified 27.6% (818/2,961; 95% CI: 26.0–29.3) of exams as *AI Negative (very high confidence)*. Of all exams, 0.7% (21/2,962; 95% CI: 0.0–1.0) were falsely classified as highconfidence negatives, corresponding to a false negative rate (FNR) of 2.0% (21/1,026; 95% CI: 0.0–2.9). Qualitative review of false negatives showed that the majority had no clinical consequence if correctly diagnosed during routine follow-up the following day, and no clearly high-risk exams were missed.

**Conclusion:** The AI system identified over one-quarter of MSK trauma radiographs as confidently negative with a very low rate of false negatives, performing on par or better than the reported standard-of-care. These results suggest potential for safe, AI-driven decision support and workflow optimization for the discharge of patients with clearly negative examinations.

## 1. Introduction

In recent years, multiple artificial intelligence (AI) decision support systems for musculoskeletal (MSK) X-ray interpretation have become commercially available [1, 3, 4, 6, 10]. These systems highlight clinical findings, such as fractures, joint dislocations, and joint effusions, using bounding boxes on radiographs. Their efficacy in improving reader accuracy is well established [1,4,5,15,18], and several studies have shown that AI can surpass reader performance in fracture detection [1, 16].

Despite these advances, AI’s impact on the clinical workflow for positive exams is limited: patients with acute findings still require evaluation and treatment by specialized personnel, leaving treatment workflows largely unchanged.

A promising and less explored application of AI lies in the identification of *negative* examinations. When an AI system can classify an X-ray as negative with very high confidence, those patients may be safely discharged, potentially reducing diagnostic bottlenecks and alleviating radiology workload. However, implementing such a workflow safely requires the false negative rate (FNR) to remain extremely low, as even a small number of missed injuries could have serious clinical consequences. Published literature reports FNRs for fracture detection in musculoskeletal (MSK) trauma radiography ranging from 1% to 7% [7, 11, 13], establishing the expected benchmark for acceptable diagnostic performance.

Radiobotics’ RBfracture, a CE-marked class IIa medical device, is the first MSK AI system designed to provide this form of decision support. It classifies relevant exams as *AI Negative (very high confidence)*, providing support for safe and efficient discharge of clearly negative X-ray MSK trauma patients. This represents a novel use of AI, integrating directly into clinical pathways to enhance efficiency rather than solely assisting in positive case detection.

**Figure 1.**
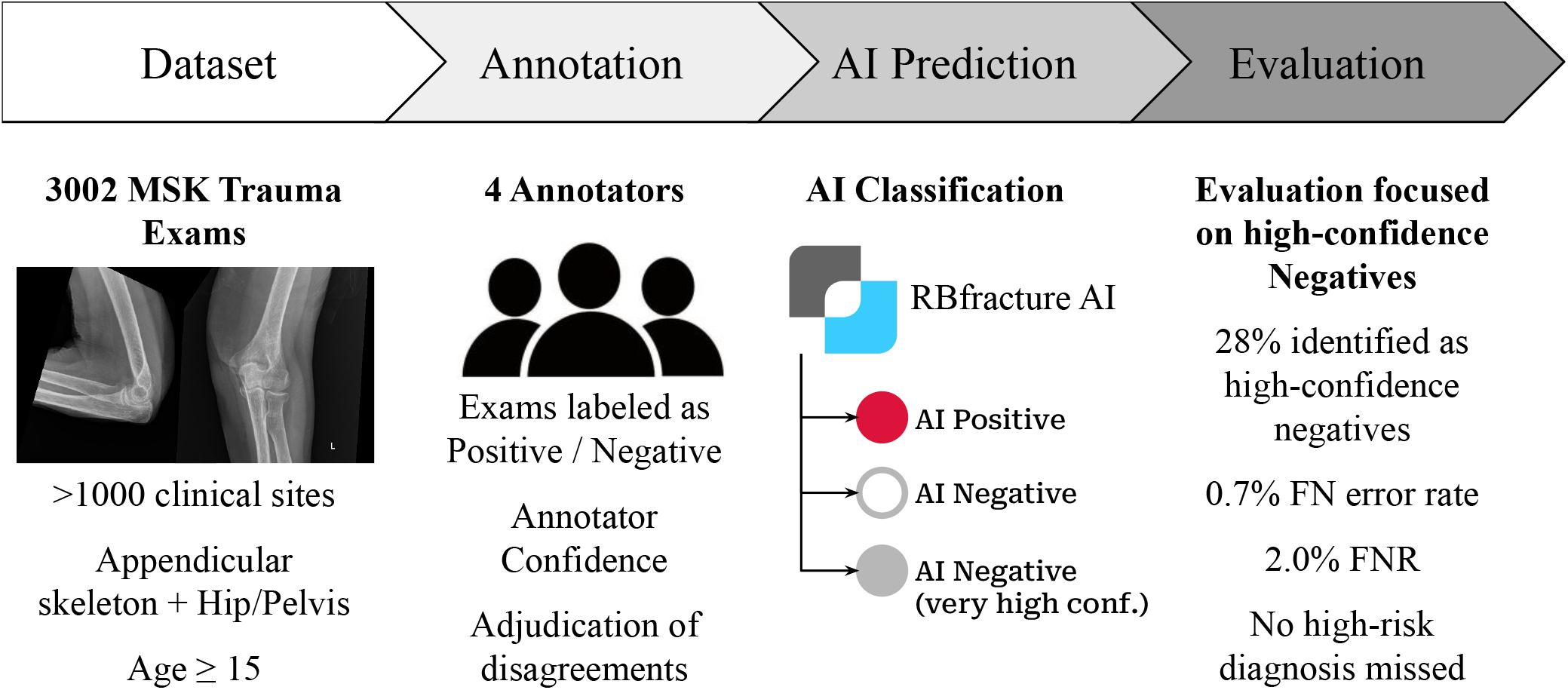
A total of 3,002 musculoskeletal (MSK) trauma exams of the appendicular skeleton including the pelvis from patients aged ≥15 years were annotated by three readers with adjudication of disagreements. Exams were analyzed with the AI system, producing three outputs (*AI Positive, AI Negative, AI Negative (very high confidence)*. The evaluation focused on high-confidence negatives, which accounted for 28% of exams with a 0.7% error rate and 2.0% false negative rate; no clearly high-risk exams were missed.

### 1.1. Study Objectives

This study evaluates the diagnostic performance of an AI system in identifying *negative* MSK trauma exams with very high confidence. We hypothesize that the system can reliably identify a substantial proportion of negative examinations while maintaining a false negative rate comparable to standard-of-care, supporting safe fast-track discharge of patients.

## 2. Methods

### 2.1. Dataset Collection

We retrospectively assembled a representative dataset of musculoskeletal (MSK) trauma examinations. For sources with available acquisition dates, we used consecutive sampling from October to November 2024; otherwise, we sampled exams at random. All data came from routine clinical practice and included radiographs, referral notes, and radiology reports. All data were anonymized before any further processing or analysis.

#### Data Sources

We obtained exams from two primary sources. The first source is a large teleradiology provider from the United States, contributing imaging from over 1,000 hospitals with substantial variation in equipment, acquisition protocols, and patient demographics. The second source is Bispebjerg and Frederiksberg Hospitals (BFH) in Copenhagen, Denmark, providing an independent validation set from a different continent. To ensure diversity while maintaining statistical balance, we weight the dataset 70% U.S and 30% Danish data.

Imaging devices and acquisition protocols vary widely across contributing sites, covering all major manufacturers.

**Inclusion and Exclusion Criteria** were established to be consistent with the intended use of the AI system and to reflect the standard-of-care. Exams were included if patients were aged ≥15 years, referred for trauma-related or acute MSK radiographic imaging, and the exam type covered one of the following regions: shoulder, upper arm, elbow, forearm, wrist, hand/fingers, hip/pelvis, upper leg, knee, lower leg, ankle, or foot/toes. Exams were excluded if they were acquired for reasons other than MSK trauma, were different exam types than those listed above, e.g., of the spine or chest or contained radiographs flagged as poor image quality by annotators.

### 2.2. Dataset Annotation

The reference standard was established by the original radiological report and four qualified annotators: three reporting radiographers with 4, 6, and 7 years of reporting experience, and one general radiologist with 13 years of experience. Annotators were selected through qualifying tests to ensure consistency and accuracy. To monitor annotation quality, 10% of all annotations were randomly reviewed by a supervisor for adherence to the annotation protocol.

#### Annotation Protocol

Each examination was initially assessed by a single annotator using a browser-based annotation platform (V7 Darwin, v7labs, London, United Kingdom). Annotators had access to the original radiographs and referral notes but were blinded to AI outputs. The primary annotation was a binary exam-level label: positive or negative. An exam was labeled positive if any acute trauma-related finding was present, including fracture, joint effusion, joint dislocation, or lipohemarthrosis. Annotators also recorded their diagnostic confidence for each positive case, classifying it as either *annotator-certain* or *annotator-equivocal*. Exams deemed insufficient for clinical interpretation were labeled *poor image quality* and excluded from further analysis.

To identify potential annotation discrepancies, each annotation was compared with its original radiology report. Reports were processed using the general large language model (LLM) GPT-4o [9], which extracted binary exam-level labels (positive or negative for trauma findings) consistent with the annotator definitions.

Cases where the initial annotation and LLM-derived report label agreed were directly accepted as the reference standard. Disagreements triggered an independent review by a second annotator, who remained blinded to the AI results. This adjudication served as the final determination and established the reference standard for those cases.

### 2.3. AI system

The evaluated AI system was RBfracture version 2.6.0, a CE-marked class IIa medical device. The AI system consists of multiple AI models designed as decision-support systems for Emergency and Radiology departments. The models automatically analyze musculoskeletal (MSK) trauma radiographs and provide structured outputs to support diagnostic workflows. It integrates with hospital PACS servers or directly with imaging modalities, producing secondary capture images. Bounding boxes highlight suspected findings in exams identified as positive, while exams identified as clearly negative are classified as *AI Negative (very high confidence)*. The system is intended for use by healthcare professionals with access to radiographic imaging and does not require specialized AI expertise.

#### Development and Training

The AI system was developed on a dataset of over 350,000 radiographs sourced from more than 1,000 radiology departments across multiple continents. Data was partitioned into a training set (80%) for model development, a tuning set (10%) for hyperparameter optimization and threshold calibration, and a test set (10%) for independent validation during development. The dataset used in this study was derived exclusively from the test set. Data from this study has not been used for training, tuning, or prior clinical evaluations of the AI system.

#### AI Outputs

The AI system generates bounding boxes to highlight and localize detected fractures, joint dislocations, joint effusions, and lipohemathrosis. In addition, it classifies the exam as a whole into one of three exam-level classes:

- *AI Positive*: indicates a suspected trauma-related finding. The finding is localized by a bounding box.
- *AI Negative*: indicates no trauma-related finding is detected.
- *AI Negative (very high confidence)*: indicates no trauma-related finding with a confidence calibrated during development to minimize false negatives and reliably identify clearly negative exams for potential workflow support.

The *AI Negative (very high confidence)* output is a newly introduced output that enables safe decision-support and workflow optimizations for negative exams with high AI confidence, and is the focus of this study.

### 2.4. Statistical Analysis

#### Outcome Definitions

For analysis, we simplified AI outputs into two categories. *Negative* corresponds to exams labeled *AI Negative (very high confidence)*, indicating that the AI is highly confident the exam is negative. *Positive* included both *AI Positive* and standard *AI Negative* outputs. This approach focused the evaluation on the highconfidence negative classifier. AI outputs were generated by processing radiographs in the DICOM format and did not include any prior data preprocessing. Exams with AI processing errors were excluded, and the total number of rejected exams was noted.

#### Classification

We evaluated agreement between the reference standard and the AI system using standard exam-level definitions:

- True Positive (TP): reference standard positive exam and correctly classified as positive by the AI system.
- False Positive (FP): reference standard negative exam but classified as positive by the AI system.
- True Negative (TN): reference standard negative exams correctly classified as *AI Negative (very high confidence)* by the AI system.
- False Negative (FN): reference standard positive exam but classified as *AI Negative (very high confidence)* by the AI system.

#### Primary Endpoint

The primary endpoint was the falsenegative rate (FNR) of the high-confidence negative classifier:

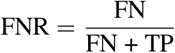

A null hypothesis for the high-confidence negative classification was pre-specified based on standard-of-care performance reported in the literature. False negative rates for fracture detection in MSK trauma radiography are reported between 1% and 7% [7, 11, 13]. In this study, we adopt an FNR of 3% as a representative benchmark for the standard-of-care and use it as the reference for formulating the hypothesis:

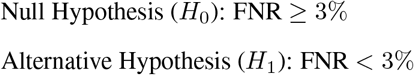

Rejection of the null hypothesis would demonstrate that the AI identified negative exams with very high confidence while maintaining a risk of missed findings less than the standard-of-care. Statistical testing was done using a one-sided exact binomial test based on the Clopper–Pearson 95% confidence interval, and an *α* = 0.05 was considered statistically significant.

All metrics were reported with 95% confidence intervals using the Clopper-Pearson method.

#### Secondary Endpoints

Secondary performance endpoints include the rate of high confidence negatives:

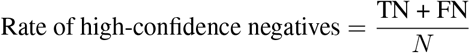

and the false negative error rate:

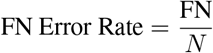

#### Subgroup and Qualitative Analyses

To assess AI robustness, the performance was stratified by annotator confidence (*annotator-certain* vs. *annotator-equivocal*), data source (U.S. vs. Denmark), patient sex (Female, Male, Unknown), and patient age (≤48, >48 years). The two age groups were separated by the median patient age. All false-negative exams were manually reviewed and categorized to evaluate potential clinical impact.

#### Sample Size

The study tests the null hypothesis that the AI system’s FNR is greater than or equal to the standard-of-care benchmark of 3% (*p*_0_ = 0.03), against the alternative hypothesis that the true FNR is lower, with an expected mean of 2% (*p*_1_ = 0.02) based on insights gained during development of the AI system. Using a significance level of *α* = 0.05 and a statistical power of 1 − *β* = 0.8, the normal approximation to the binomial distribution indicates that a minimum of 753 positive exams is required to detect this difference. Since the strategy was to sample the cases consecutively to reflect real-world performance a minimum sample size of 2,151 was needed assuming the prevalence was 35% (753/0.35). To account for uncertainties about the actual performance and actual prevalence the desired sample size was chosen to be N=3,000 consecutively sampled patients.

## 3. Results

### 3.1. Study Population

A total of 3,002 pseudo-consecutive musculoskeletal (MSK) trauma radiographic examinations were retrospectively sampled. Exams from the U.S. were sampled consecutively from September to October 2024, restricted to referrals explicitly indicating suspected trauma. For the Danish data source, random sampling was applied due to missing timestamps. Inclusion was limited to patients aged ≥15 years and to exam types specified in the study criteria. Filtering was based on DICOM metadata, which was not always reliable, leading to some excluded exam types being rejected for processing later.

All exams were annotated by the first annotator according to the annotation protocol. In 585 of 3,002 exams (19.5%), potential discrepancies between the annotator and the original radiographic report were flagged for adjudication by the LLM. The adjudicator overruled the annotations in favour of the radiology report in 115 of the 585 flagged exams (3.8% of the study population).

Twenty-five exams were excluded during reference standard establishment: four due to poor image quality and 21 due to excluded exam types (20 chest, 1 skull). This yielded 2,977 exams for AI analysis.

The AI system processed all eligible exams. Fifteen exams were excluded due to AI processing errors (`NoSupportedInput`; n=9, `OutsideIntendedUse`; n=6). The final study population consisted of 2,962 exams (see Figure 2).

**Figure 2.**
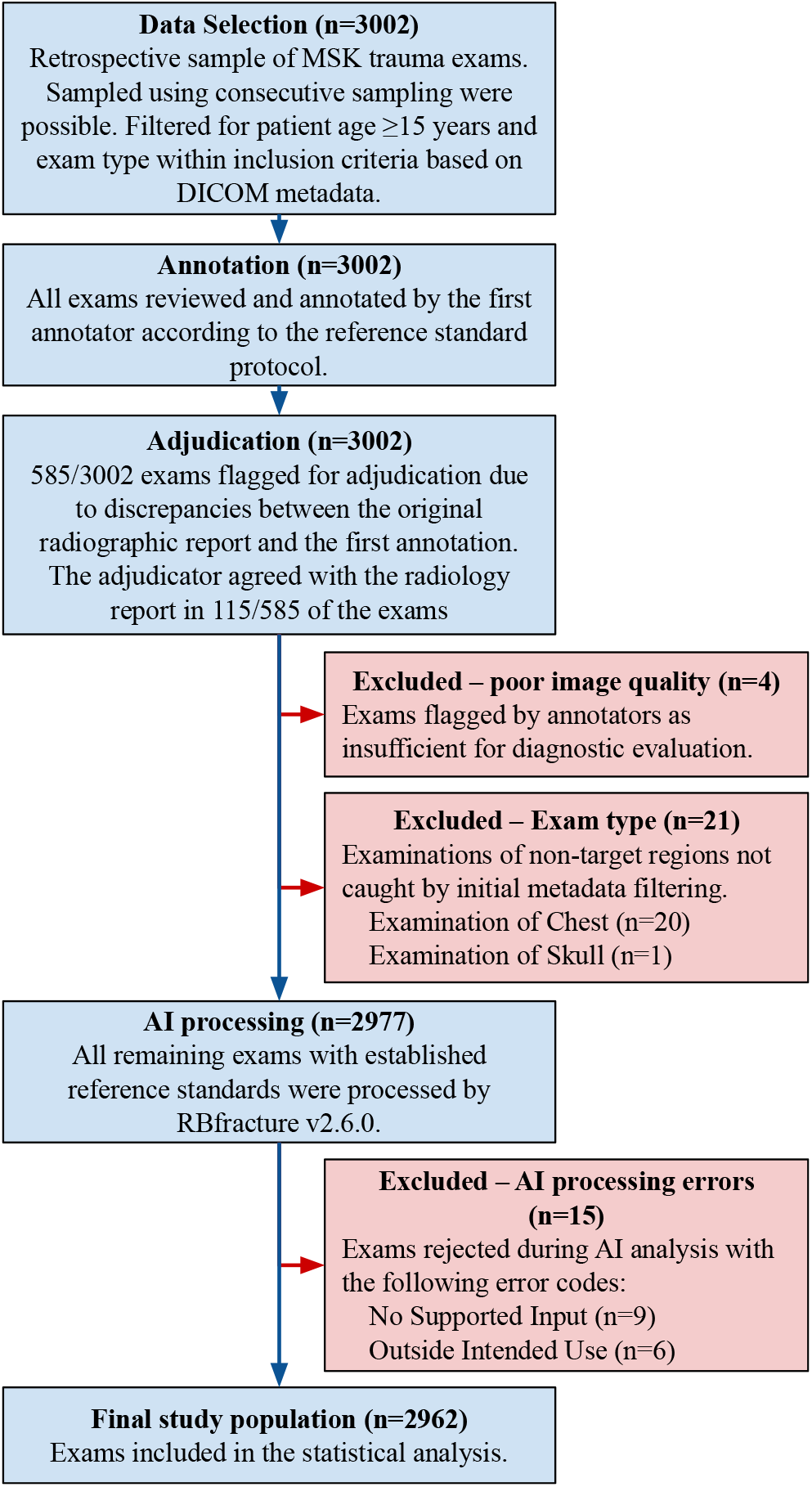
Flow of exams from initial selection to final study population. The diagram shows annotation, adjudication, exclusions, AI processing, and the number of exams included in the analysis.

Of these, the median patient age was 48 years (range 15–103; 25% quartiles 29, 69 years). The patient’s sex is known for 30.6% of exams, and anonymized for the remainder. Out of the exams with known sex, 50.8% of patients were female and 49.2% were male. All exam types specified in the inclusion criteria were represented, with distributions consistent with those typically observed in trauma clinics (Table 1).

**Table 1.** Distribution of radiographic exams by type, patient age, data source (U.S. and Danish), and patient sex. The exam type “Unknown” refers to exams for which the type could not be automatically inferred from DICOM metadata. Patient Sexes have been anonymized on the U.S. data source.

| Exam type | No. | Age (median, quartiles) | U.S. | Danish | Female | Male | Unknown Sex |
| --- | --- | --- | --- | --- | --- | --- | --- |
| Foot & Toes | 409 | 39 (26, 56) | 249 | 160 | 74 | 86 | 249 |
| Ankle | 416 | 37 (26, 55) | 228 | 188 | 100 | 88 | 228 |
| Lower Leg | 143 | 50 (32, 66) | 97 | 46 | 23 | 23 | 97 |
| Knee | 405 | 51 (30, 70) | 222 | 183 | 99 | 84 | 222 |
| Upper Leg | 85 | 73 (52, 80) | 57 | 28 | 17 | 11 | 57 |
| Hip & Pelvis | 264 | 74 (61, 80) | 165 | 99 | 54 | 45 | 165 |
| Hand & Fingers | 412 | 39 (26, 59) | 321 | 91 | 28 | 63 | 321 |
| Wrist | 227 | 49 (28, 68) | 183 | 44 | 28 | 16 | 183 |
| Forearm | 88 | 41 (30, 68) | 82 | 6 | 3 | 3 | 82 |
| Elbow | 148 | 52 (30, 66) | 129 | 19 | 10 | 9 | 129 |
| Upper Arm | 66 | 67 (45, 80) | 62 | 4 | 3 | 1 | 62 |
| Shoulder | 248 | 52 (33, 72) | 215 | 33 | 18 | 15 | 215 |
| Unknown | 51 | 64 (44, 79) | 50 | 1 | 1 | 0 | 50 |
| Total | 2962 | 48 (29, 69) | 2060 | 902 | 458 | 444 | 2060 |

The reference standard identified 1,026 positive exams, corresponding to a prevalence of 34.6%. In 104 exams (10.1% of positives), annotators expressed diagnostic uncertainty with the *annotator-equivocal* tag, reflecting the inherent difficulty of some trauma presentations.

### 3.2. Test results

The AI system was evaluated on 2,962 musculoskeletal trauma examinations. A cross-tabulation of the predictions and reference standard is provided in Table 2, and performance is summarized in Table 3. The system produced 1,135 *AI Positive* predictions, 1,009 *AI Negative* predictions, and 818 *AI Negative (very high confidence)* predictions.

**Table 2.** Cross-tabulation of AI outputs against the reference standard. Positive exams in the reference standard are stratified by *annotator-certain* and *annotator-equivocal*. Negative exams are also shown.

| AI Output | Reference standard |  |  | Total |
| --- | --- | --- | --- | --- |
|  | Positive (certain) | Positive (equivocal) | Negative |  |
| AI Positive | 865 | 55 | 215 | 1,135 |
| AI Negative | 50 | 35 | 924 | 1,009 |
| AI Negative (very high confidence) | 7 | 14 | 797 | 818 |
| Total | 922 | 104 | 1,936 | 2,962 |

**Table 3.** Diagnostic performance of the *AI Negative (very high confidence)* AI output class. Performance was reported for all 2,962 exams and stratified by annotator confidence, data source, patient sex, and patient age. Metrics include false-negative rate (FNR), false-negative (FN) error rate, and the proportion of exams classified as high-confidence negative by the AI, each with 95% Clopper–Pearson exact confidence intervals. ^1^ The number of False Positives appears high, as we count Reference standard Negative exams classified as *AI Negative* as false positives in this evaluation.

| Subgroup | FNR (%) | FN error rate (%) | Rate of high conf. negatives (%) | TP | TN | FP<sup>1</sup> | FN |
| --- | --- | --- | --- | --- | --- | --- | --- |
| All Exams | 2.0 (0–2.9) | 0.7 (0–1.0) | 27.6 (26.0–29.3) | 1005 | 797 | 1139 | 21 |
| *annotator-certain* Exams | 0.8 (0–1.4) | 0.2 (0–0.5) | 28.1 (26.5–29.8) | 915 | 797 | 1139 | 7 |
| U.S. Exams | 1.6 (0–2.7) | 0.5 (0–0.8) | 28.3 (26.4–30.3) | 615 | 574 | 861 | 10 |
| Danish Exams | 2.7 (0–4.5) | 1.2 (0–2.0) | 25.9 (23.1–28.9) | 390 | 223 | 278 | 11 |
| Female Patients | 2.5 (0–5.1) | 1.1 (0–2.3) | 27.3 (23.3–31.6) | 198 | 120 | 135 | 5 |
| Male Patients | 3.0 (0–5.9) | 1.4 (0–2.6) | 24.5 (20.6–28.8) | 192 | 103 | 143 | 6 |
| Unknown Sex | 1.6 (0–2.7) | 0.5 (0–0.8) | 28.3 (26.4–30.3) | 615 | 574 | 861 | 10 |
| Age ≤ 48 years | 2.2 (0–3.7) | 0.7 (0–1.1) | 36.7 (34.3–39.2) | 442 | 537 | 500 | 10 |
| Age > 48 years | 1.9 (0–3.2) | 0.7 (0–1.2) | 18.4 (16.4–20.5) | 563 | 260 | 639 | 11 |

The false negative rate was 2.0% (21/1,026; 95% CI: 0.0–2.9%), below the pre-specified benchmark of 3%. As the upper bound of the 95% confidence interval did not exceed 3%, the null hypothesis (*H*_0_: FNR >3%) was rejected, indicating that the AI system achieved performance comparable to or better than the standard-of-care. High-confidence negatives accounted for 27.6% (818/2,961; 95% CI: 26.0–29.3%) of all examinations.

#### Impact of annotator certainty

Performance varied with the certainty of the reference standard. Restricting the analysis to *annotator-certain* exams reduced the FNR to 0.8% (7/922; 95% CI: 0.0–1.4%) and the false-negative error rate to 0.2% (7/2,858; 95% CI: 0.0–0.5%) (Table 3). This indicated that AI performance was highest when the diagnostic certainty of the ground truth was non-equivocal.

#### Impact of data source

Performance was similar across regions. The AI system achieved an FNR of 1.6% (10/625; 95% CI: 0.0–2.7%) on U.S. examinations and 2.7% (11/401; 95% CI: 0.0–4.5%) on Danish examinations. The proportion of exams classified as *AI Negative (very high confidence)* was slightly higher for U.S. cases at 28.3% (584/2060; 95% CI: 26.4–30.3%) compared to 25.9% (234/902; 95% CI: 23.1–28.9%) for Danish cases. Although the U.S. data showed marginally better performance, the overlapping confidence intervals indicate that the differences were minor (Table 3).

#### Impact of patient sex

Performance was consistent across sexes, with an FNR of 2.5% (5/203; 95% CI: 0.0– 5.1%) for females and 3.0% (6/198; 95% CI: 0.0–5.9%) for males. The rate of high confidence negative predictions was also similar, at 27.3% (125/458; 95% CI: 23.3–31.6%) for females and 24.5% (109/444; 95% CI: 20.6–28.8%) for males (Table 3).

#### Impact of patient age

Large differences in the rate of *AI Negative (very high confidence)* predictions were observed across age groups. The younger half of the patient population had a prediction rate of 36.7% (547/1489; 95% CI: 34.3–39.2%) while the older half had a rate of 18.4% (271/1473; 95% CI: 26.4–20.5%). The FNR was similar across both age groups, at 2.2% (10/452; 95% CI: 0.0– 3.7%) for patients aged ≤48 years, and 1.9% (11/574; 95% CI: 0.0–3.2%) for patients aged *>*48 years. (Table 3).

#### Patient flow and exam distribution

The distribution of exams across reference categories and AI outputs is shown in Figure 3. Most reference-standard positives were classified as *AI Positive* (n=920), with fewer assigned to *AI Negative* (n=85) or *AI Negative (very high confidence)* (n=21). Within the high-confidence negative group, false negatives represented only 21 of 818 exams (2.6%), indicating that missed findings comprised a very small fraction of confidently negative classifications.

**Figure 3.**
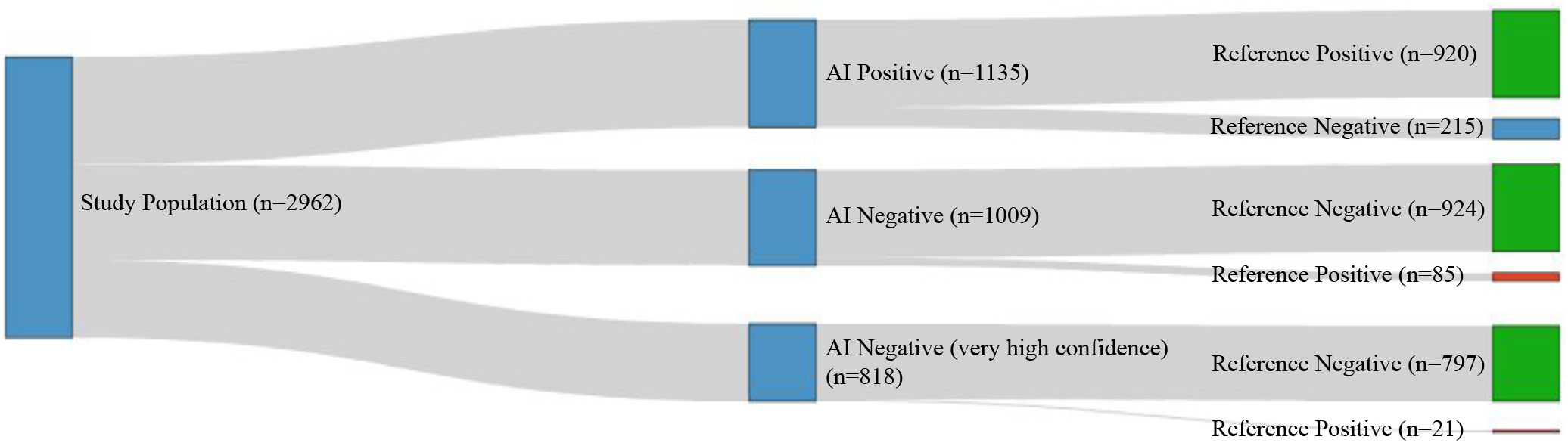
Patient flow from study population (column 1), to AI outputs (column 2), to reference standard annotation (column 3).

#### Qualitative evaluation of false negatives

All 21 false negative exams were reviewed by a reporting radiographer to assess potential clinical consequences of delayed diagnosis resulting from the incorrect AI prediction. Two example exams are shown in Figure 4. Of all false negatives, 14 were fractures, 5 were joint effusions, and 2 were joint dislocations. The fractures and effusions were judged to have no clinical consequence if diagnosed during routine review on the following day. In contrast, the two dislocations were considered potentially clinically consequential. Both dislocations had been labeled as *annotator-equivocal* in the reference standard, reflecting their inherent diagnostic difficulty (Table 4). A detailed description of all exams is given in the appendix.

**Table 4.** Qualitative review of the 21 false-negative exams, representing 0.7% of the study population. A reporting radiographer categorized exams based on clinical impact if diagnosis were delayed until the following day. A detailed description of all exams is given in the appendix.

| False Negatives |  |
| --- | --- |
| Categorization | Details |
| Pathology | 14x Fracture |
|  | 5x Joint effusion |
|  | 2x Joint dislocation |
| Annotator certainty | 7x *annotator-certain* |
|  | 14x *annotator-equivocal* |
| Clinical impact | 19x no clinical consequence if diagnosed the following day |
|  | 2x potentially clinically consequential even if diagnosed the following day |

**Table 5.** Qualitative evaluation of the 21 False Negative exams. A reporting radiographer provided pathology localization and descriptions, classified each finding in the AO/OTA Fracture and Dislocation Classification system [14], and assessed the clinical impact if the pathology were initially misdiagnosed due to the AI classification, but identified during routine review on the following day.

| No. | Pathology | Annotator Certainty | AO Class | Exam Type | Localization | Description |
| --- | --- | --- | --- | --- | --- | --- |
| 1 | Joint Dislocation | Equivocal | 80.D | Foot & Toes | Intermetatarsal | An increased distance between the base of the 1st and 2nd metatarsal bone cannot be excluded, Lisfranc |
| 2 | Joint Dislocation | Equivocal | 10.A | Shoulder | Humeral Joint | Suspected posterior shoulder joint dislocation. Disagreement between radiologist (reporter and annotator) |
| 3 | Joint Effusion | Certain | – | Knee | Knee Suprapatellar Recess | Knee joint effusion in the suprapatellar recess |
| 4 | Joint Effusion | Certain | – | Knee | Knee Suprapatellar Recess | Knee joint effusion in the suprapatellar recess cannot be ruled out |
| 5 | Joint Effusion | Equivocal | – | Knee | Knee Suprapatellar Recess | Knee joint effusion in the suprapatellar recess cannot be ruled out |
| 6 | Joint Effusion | Certain | – | Knee | Knee Suprapatellar Recess | Knee joint effusion in the suprapatellar recess |
| 7 | Joint Effusion | Certain | – | Knee | Knee Suprapatellar Recess | Knee joint effusion in the suprapatellar recess cannot be ruled out |
| 8 | Fracture | Certain | 88.4.2.1B | Foot & Toes | 4th Toe, Phalanx Media | 4th phalanges media, proximal segment, partial intraarticular |
| 9 | Fracture | Equivocal | 85.1.A | Foot & Toes | Cuneiforme Media | Undisplaced fracture in the medial part of cuneiforme media cannot be ruled out |
| 10 | Fracture | Equivocal | 4F3A | Ankle | Lateral Malleolus | Simple, non-displaced, intraarticular, oblique fracture in the lateral malleolus cannot be ruled out |
| 11 | Fracture | Equivocal | 2R3B | Hand & Fingers | Distal Radius | Larger non-displaced avulsion in the rim of the distal radius cannot be ruled out |
| 12 | Fracture | Equivocal | 85...A | Foot & Toes | Cuneiforme | Suspected small, non-displaced avulsion from a cuneiforme |
| 13 | Fracture | Equivocal | 44.A.3 | Ankle | Distal Tibia and Malleolus | Fracture in posterior distal tibia as well as non-displaced fracture of the medial malleolus. Doubtful or invisible in the single image available |
| 14 | Fracture | Certain | – | Ankle | Tarsal | Suspected fracture in the midfoot, questionable from which tarsal. Not reported. |
| 15 | Fracture | Equivocal | 76.2 | Wrist | Triquetrum | Suspected avulsion of the triquetrum |
| 16 | Fracture | Certain | 88.1.2.3C | Foot & Toes | 1st Toe, Distal Phalanx | Non-displaced, intraarticular fracture in the 1st distal phalanges |
| 17 | Fracture | Equivocal | 44A1.2 | Foot & Toes | Lateral Malleolus | Avulsion in the distal tip of the lateral malleolus. Reported from the ankle exam, not visible in this foot exam |
| 18 | Fracture | Equivocal | 4F2A | Lower Leg | Distal Fibula Diaphysis | Questionable non-displaced oblique fracture in the distal segment of the fibula diaphysis |
| 19 | Fracture | Certain | 88.1.2.3B | Foot & Toes | 1st Toe, Distal Phalanx | Non-displaced, intraarticular fracture in the rim of the 1st distal phalanges |
| 20 | Fracture | Certain | 87.5.2.A | Ankle | 5th Metatarsal Diaphysis | Oblique fracture in the diaphysis of the 5th metatarsal. Reported on the foot exam; in this ankle exam it is barely visible |
| 21 | Fracture | Equivocal | 82 | Ankle | Calcaneus | Non-displaced avulsion of the calcaneus anterior process |

**Figure 4.**
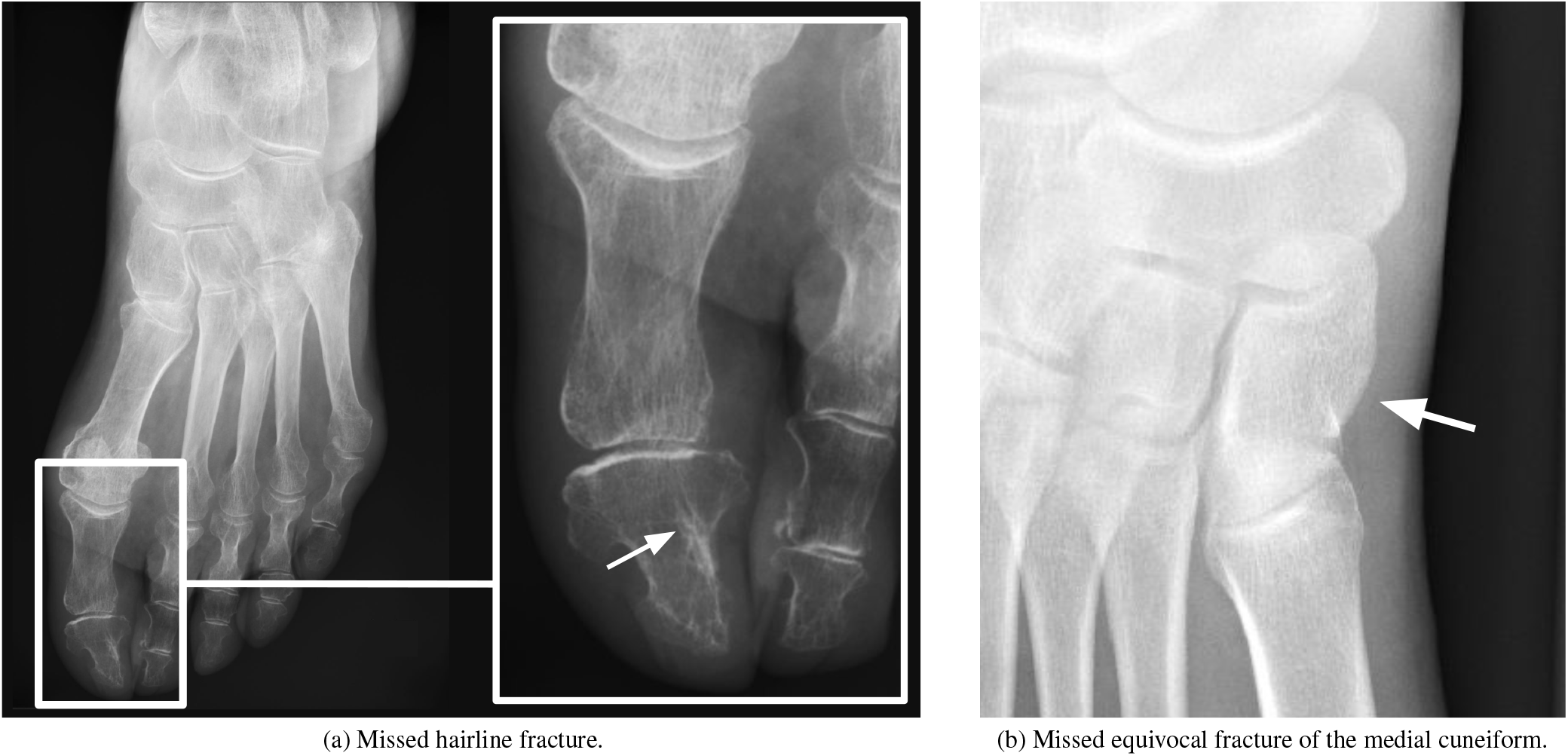
Examples of False Negatives. A white arrow highlights the reference standard annotation. Both exams have been judged as not clinically consequential if initially misdiagnosed due to the AI classification, but identified during routine review on the following day. **Left**: A hairline fracture in the distal phalanges of the first toe. Annotated as reference standard positive (*annotator-certain*), but classified as *AI Negative (very high confidence)* by the AI system. **Right**: A non-displaced fracture in the medial part of medial cuneiform cannot be ruled out. Annotated as *annotator-equivocal*, but classified as *AI Negative (very high confidence)* by the AI system.

## 4. Discussion

This study evaluated the diagnostic performance of the AI system RBfracture for musculoskeletal trauma radiography, with a focus on the identification of clearly negative exams. The study met its primary endpoint, demonstrating that the AI system’s high-confidence negative classifier achieved a false negative rate below the 3% benchmark, representing standard-of-care performance. High-confidence negative predictions accounted for more than one quarter of all examinations, demonstrating that the AI identified a meaningful subset of patients as clearly negative.

When analysis was limited to exams confidently labeled by annotators, the FNR decreased to 0.8%, suggesting that the AI achieved its highest accuracy when the reference standard reflected clear diagnostic certainty.

The AI system performed consistently across patient sexes and data sources, producing overlapping confidence intervals in all performance metrics.

A substantial difference was observed in the rate of high-confidence negative AI predictions across age groups: patients aged ≤48 years had twice the proportion of high-confidence negative AI predictions compared with patients >48 years. This pattern aligns with clinical experience. The exclusion of findings is often more straightforward in younger individuals, whose radiographs show fewer confounding features. In contrast, older patients often present with age-related degenerative changes, reduced bone density, prior injuries, or anatomical remodeling, all of which complicate image interpretation. These factors likely reduce the certainty with which both clinicians and the AI system can rule out fractures in older patients, explaining the lower rate of high-confidence negative predictions in this group.

A qualitative review of the 21 false-negative exams showed that the majority (19 of 21) were fractures or joint effusions with no clinical consequence if diagnosed during routine review on the following day. Two joint dislocations were judged as potentially clinically consequential if diagnosis were delayed; both, however, had been labeled as equivocal in the reference standard, reflecting genuine diagnostic difficulty. No clearly high-risk or clinically urgent findings were missed in the high-confidence group.

### Comparison to standard-of-care and other AI systems

Reported standard-of-care false negative rates (FNR) in musculoskeletal trauma imaging range from 1% to 7% [7, 11, 13], with a benchmark of 3% used in this study. The AI system achieved an FNR of 2.0%, which surpassed the benchmark. Prior studies on commercial fracture detection AI systems report FNRs of 3%–14% [2, 8, 12, 17, 18], performing much worse at identifying clearly negative exams.

### Limitations

This study has several limitations. First, as a standalone evaluation, it did not assess the impact of the AI system on clinical workflows, reader performance, or patient outcomes. Further prospective studies are required to evaluate how the system integrates into real-world radiology practice. Second, the dataset was sourced exclusively from the U.S. and Denmark, which may limit generalizability to settings with different imaging protocols, patient populations, or disease prevalence. Third, the standard-of-care baseline (3% FNR) was derived from published literature rather than measured directly on the study dataset. While the standard-of-care performance is assessed across all exams, the AI system was evaluated only on a subset where it was highly confident. Fourth, although the training and evaluation datasets did not overlap at the patient level, both were drawn from the same overall data sources, introducing potential bias. Finally, the analysis was restricted to acute trauma-related findings (fracture, joint dislocation, joint effusion, and lipohemarthrosis); non-trauma or chronic findings were not assessed.

### Clinical implications

Despite these limitations, the findings support a clinically relevant use case: AI-assisted discharge decisions of patients confidently classified as negative. The very low FNR observed suggests that the AI could safely reduce radiology workload by automatically excluding a meaningful fraction of exams without compromising patient safety. Future work will evaluate this potential in prospective clinical studies, including its impact on turnaround times, radiologist efficiency, and patient flow.

## 5. Conclusion

The negative classification with *very high confidence*, introduced as a novel feature in RBfracture v2.6.0, demonstrated robust diagnostic performance in musculoskeletal trauma imaging, achieving a false negative rate below reported standard-of-care benchmarks in exams classified by the AI as confidently negative. Qualitative review of errors confirmed that no definitive high-risk findings were missed. These results indicate that the system can safely identify a substantial proportion of examinations as negative, with potential to improve efficiency in trauma radiology workflows. Future research will evaluate its impact in prospective, realworld settings and explore broader generalizability across healthcare systems.

### Ethics Statement

The dataset consisted of retrospectively collected, fully anonymized musculoskeletal trauma radiographs obtained from two sources: a U.S.-based teleradiology company and the Department of Radiology at Bispebjerg and Frederiksberg Hospital, Denmark. Collaboration agreements were established between the data providers and Radiobotics. Inclusion of anonymised data was approved, and patient consent was waived by the Institutional Review Board at WCG Clinical (IRB Tracking Number: 20254498) and Danish Patient Safety Authorities (Case number: 3-3013-3040/1). AI was used to assist in text editing and programming.

### Funding & Commercial interests

All authors are employees at Radiobotics, the company developing and marketing the evaluated AI system RBfracture. The study did not receive any funding.

### Author Contributions

**Steffen Czolbe:** Methodology, Software, Visualization, Writing – Original Draft; **Jonas Christophersen:** Software, Validation, Data Curation, Writing – Review & Editing; **Rikke Bachmann:** Investigation, Data Curation, Validation, Writing – Review & Editing; **Janitha Mudannayake:** Validation; **Andreas Nexmann:** Software, Data Curation; **Fabizio Perria:** Software; **Nikolaus Knauer:** Project administration; **Pavel Lisouski:** Conceptualization; **Michael Lundemann:** Conceptualization, Writing – Review & Editing.

## Data Availability

All data produced in the analysis and results present study are available upon reasonable request to the authors. Raw imaging data is not available.

## Appendix

**Qualitative evaluation of False Negative exams**

